# Evaluation of Junior Doctors’ Knowledge of Corneal Donation and the New Opt-Out System in England

**DOI:** 10.1101/2021.03.04.21252895

**Authors:** Bhavesh Gopal, Owuraku Asiedu Titi-Lartey, Princeton Fernandes, Nur-Emel Noubani, Elizabeth Blatherwick, Dalia G. Said, Harminder S. Dua, Darren Shu Jeng Ting

## Abstract

**Objectives:** To evaluate the knowledge of corneal donation and the new opt-out system among junior doctors in the East Midlands, UK.

**Methods:** This was a cross-sectional study performed during June-September 2020. A 26-item questionnaire-based survey was disseminated to all 340 junior doctors working in the East Midlands, UK. Relevant data, including participants background, knowledge of corneal donation and the new opt-out system introduced in England, were analysed.

**Results:** A total of 143 responses were received (response rate=42.1%). Nineteen (13.3%) junior doctors had previously discussed about corneal donation. The majority (100, 69.9%) of them perceived the importance of obtaining consent for corneal donation as junior doctors, but only 24 (16.8%) felt comfortable in discussing corneal donation. The knowledge of corneal donation was low, with a mean correct response rate of 33.3+/-20.8%. Only 28 (19.6%) doctors were aware of the 24-hour death-to-enucleation time limit. The majority (116, 81.1%) of doctors would consider certifying a death on the ward quicker if they knew it could potentially compromise the quality of corneas. Most (103, 72%) doctors were aware of the new opt-out system but only 56 (39.2%) doctors correctly stated that donation can only proceed with family consent.

**Conclusion:** Junior doctors working at the frontline services serve as valuable members in contributing to the process of obtaining consent for organ/tissue donation. Our study highlights the lack of knowledge of corneal donation and the opt-out system amongst junior doctors in the UK. Targeted postgraduate training during the induction process may potentially enhance the donation rate.

## INTRODUCTION

Corneal opacity represents the 5^th^ leading cause of blindness and visual impairment globally, affecting around 6 million of the population.^1^ Corneal transplantation serves as the mainstay of treatment in restoring vision in patients affected by corneal opacity.^2^ It is the most commonly performed transplantation worldwide, though the success has been persistently challenged by the global shortage of donor cornea.^2^

To date, a wide range of initiatives and advancement, including public campaigns to increase awareness, introduction of telephone consent, refinement in the donation-transplantation pathway and improvement in surgical techniques,^3-6^ have been implemented to improve corneal donation and utility of donor corneal tissues. On 20^th^ May 2020, England has implemented an opt-out system, also known as the Max and Keira’s Law, with an aim to improve the rate of organ/tissue donation, joining countries such as Spain, France and Italy, and many others. Under the new, soft opt-out system, all adults in England are assumed to be willing organ/tissue donors unless they have registered their intent otherwise. However, the process of eye donation remains largely unchanged as consent from the family members is still required before retrieval can proceed.

Junior doctors at frontline services, particularly those who work in intensive care, oncology and palliative care units, may serve as valuable members to the multi-disciplinary team in contributing to the process of organ/tissue donations. Nonetheless, the knowledge of corneal donation and the new opt-out system among junior doctors in the UK has not been explored. Our study aimed to evaluate the knowledge of corneal donation and the opt-out system among the junior doctors in East Midlands, UK.

## MATERIALS AND METHODS

This was a cross-sectional study performed between 28 June 2020 and 29 September 2020. A 26-item questionnaire-based online survey was distributed to the junior doctors / house officers (<3 years of medical practice) who were working in the East Midlands, UK. The questionnaire was composed of 26 questions, which evaluated the background and ophthalmology training of participants (6 items), knowledge of corneal (and tissue) donation (13 items) and the new opt-out system (2 items), views and experience in obtaining consent for corneal donation (4 items), and views on certifying death (1 item). The detail of the questionnaire is provided in **Supplementary Figure 1**. The questionnaire was devised based on the previous studies^7,8^ and the suitability and contraindications to corneal donation set by the guideline of the National Health Service Blood and Transplant (NHSBT; https://www.transfusionguidelines.org/red-book/chapter-21-tissue-banking-tissue-retrieval-and-processing/21-2-retrieval). Ethics approval was obtained from the Research Ethic Committee at the University of Nottingham, UK, prior to the conduct of study (Reference: FMHS 45-0720). The study was conducted in accordance with the tenets of the Declaration of Helsinki.

## RESULTS

### Characteristics and experience of junior doctors

Of all 340 junior doctors, 143 survey responses were received (42.1%, response rate). The amount of previous undergraduate teaching on ophthalmology was 11.4±12.1 days (range, 0-90 days; **Table 1**). Only 24 (16.8%) junior doctors had undergone an ophthalmology training rotation during their Foundation Year training. The mean number of discussions of corneal donation held between the junior doctors and the potential donors’ family members was 0.2±0.6 (range, 0-4), with the majority (124, 86.7%) of junior doctors having never held any discussion on corneal donation. However, 100 (69.9%) of them felt that it was important to know how to obtain consent for corneal donation as a junior doctor but 119 (83.2%) were uncomfortable in discussing corneal donation with the family members.

**Table 1.** Background of ophthalmology training and intent for corneal donation among the junior doctors.

| Parameters | Total N = 143<br>N (%) |
| --- | --- |
| Level of training |  |
| Foundation Year 1 | 83 (58.0) |
| Foundation Year 2 | 50 (35.0) |
| Other | 10 (7.0) |
| Amount of undergraduate ophthalmology teaching, days |  |
| 0 | 5 (3.5) |
| 1-7 | 55 (38.5) |
| 8-14 | 65 (45.5) |
| 15-30 | 11 (7.7) |
| >30 | 7 (4.9) |
| Any ophthalmology rotation during Foundation Year (FY) training |  |
| Yes | 24 (16.8) |
| No | 119 (83.2) |
| Willingness to donate corneas |  |
| Yes | 87 (60.8) |
| No | 15 (10.5) |
| Uncertain | 41 (28.7) |
| Number of previous discussions with potential donors or their family members on corneal donation |  |
| 0 | 124 (86.7) |
| 1-2 | 17 (11.9) |
| >2 | 2 (1.4) |
| Felt that it was important to know how to obtain consent for corneal donation |  |
| Yes | 100 (69.9) |
| No | 26 (18.2) |
| Maybe | 17 (11.9) |
| Comfortable in discussing corneal donation |  |
| Very or somewhat comfortable | 24 (16.8) |
| Neutral | 45 (31.5) |
| Very or somewhat uncomfortable | 74 (51.7) |
| Felt that undergraduate teaching was sufficient for discussing corneal donation |  |
| Yes | 6 (4.2) |
| No | 137 (95.8) |
| Sources of corneal donation information |  |
| Undergraduate teaching | 43 (30.1) |
| FY training | 18 (12.6) |
| Internet | 71 (49.7) |
| Social media | 23 (16.1) |
| Health professionals | 32 (22.4) |
| Friends | 21 (14.7) |

### Knowledge of corneal (and tissue) donation and the new opt-out system

The knowledge of corneal (and tissue) donation, including 3 items on ocular conditions and 10 items on general health or systemic conditions, among the junior doctors is summarised in **Figure 1**. Overall, the mean correct response rate was 31.9±20.4% (range, 0-76.9%).

**Figure 1.**
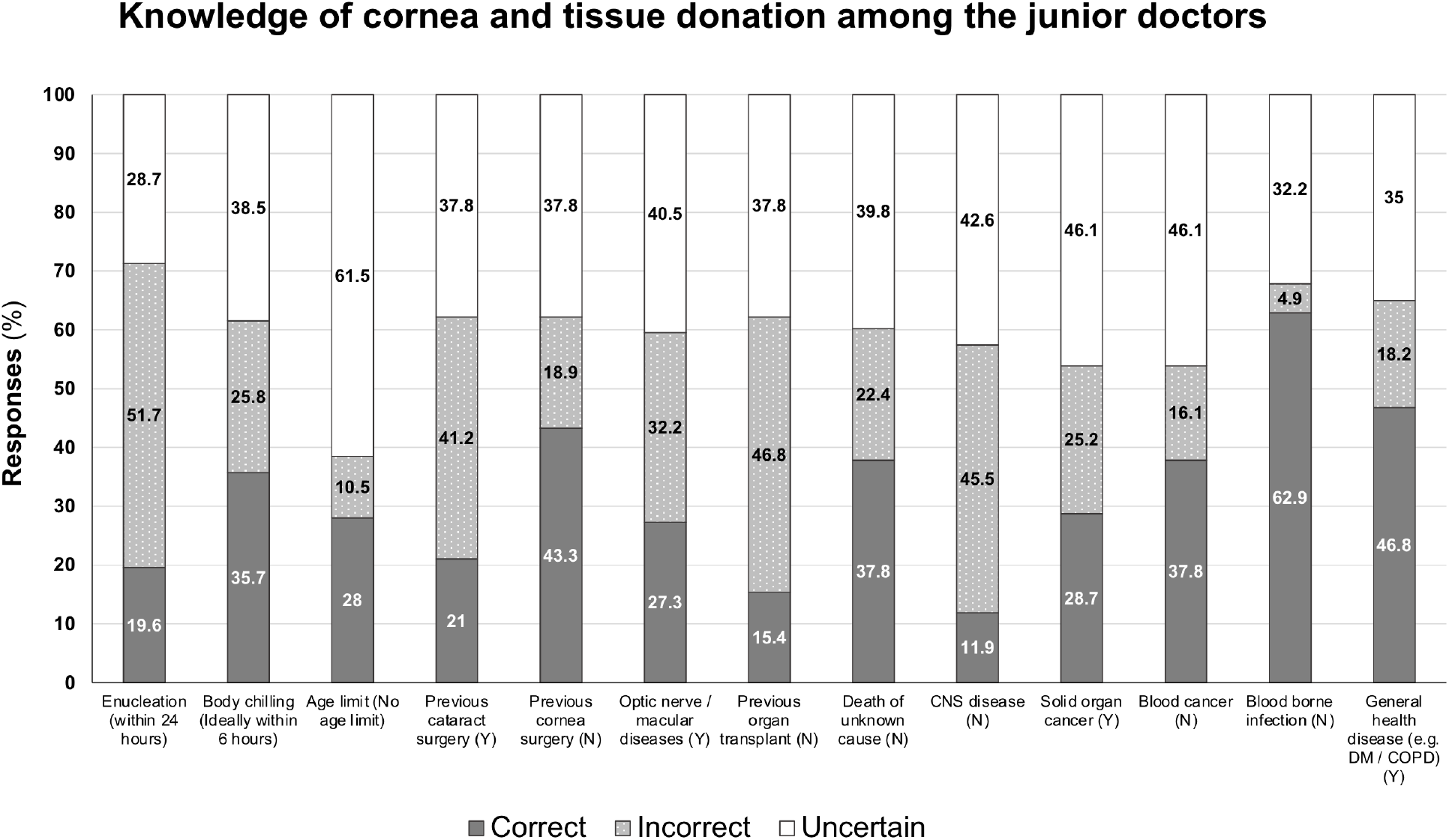
A summary of the knowledge of corneal and tissue donation amongst junior doctors in the UK. Columns 1-3 and 7-13 are related to general health or systemic conditions whereas columns 4-6 are related to ocular conditions. For each question, the correct response is provided in bracket (Y = Suitable for donation; N = Not suitable for donation). The answers provided by the respondents are divided into either correct, incorrect or uncertain responses. CNS = Central nervous system; DM = Diabetes mellitus; COPD = Chronic obstructive pulmonary disease

Only 28 (19.6%) and 51 (35.7%) junior doctors were correct about the 24-hour death-to-enucleation and the ideal 6-hour death-to-body chilling time limit (for tissue donation). Only 40 (28.0%) junior doctors were correct that there was no age limit for corneal donation. The knowledge of corneal donation was not influenced by the level of the junior doctors (p=0.36), amount of undergraduate ophthalmology teaching (p=0.19), previous ophthalmology rotation (p=0.71), previous experience in discussing corneal donation (p=0.73), and willingness to donate their own corneas (p=0.16; **Supplementary Table 1**). The majority (116, 81.1%) of junior doctors would consider certifying a death on the ward quicker if they knew it could compromise the quality of corneas.

The majority (103, 72%) of junior doctors were aware of the recently introduced opt-out system in England. In the event of family members refusing to donate organs/tissues of the deceased patient, only 56 (39.2%) junior doctors correctly stated that the organs and tissues cannot be retrieved.

## DISCUSSION

To the best of our knowledge, this represents the first study examining the views and knowledge of corneal donation and the new opt-out system among junior doctors in the UK. Currently, in the UK, consent for corneal donation is primarily obtained by a team of well-trained, specialised nurses from the National Referral Centre (NRC) embedded within the NHSBT. This process takes place as soon as the death has been certified and notified to the NRC. Although junior doctors working at frontline services are not expected to obtain consent for corneal donation in the UK, they are the key multi-disciplinary members who have daily contact with the patients and potential organ/tissue donors and may therefore play an important role in the process of organ and tissue donations (**see Supplementary Figure 2** for an example of the eye donation process in England). In addition, junior doctors are usually the key members in discussing the advance directives such as “Do not attempt cardiopulmonary resuscitation (DNACPR)” with the family members. Therefore a successful relationship of trust has already been built throughout the process of care, which could improve the conversion rate of corneal donation.^9^ Our study showed that around 13% of the junior doctors had been involved in the discussion of corneal donation with the family members, and this figure is likely to increase under the new opt-out system. Studies have shown that prior knowledge of corneal donation serves as an important factor in influencing the willingness of donating the corneas.^10^ Therefore, if an earlier discussion on corneal donation can be held between the junior doctors and potential donors’ family members, the chances of corneal donation can be potentially improved when it comes to the stage of formal consenting by the NRC.

The NHSBT has set a 24-hour cut-off interval between death and retrieval of donor corneas and the body should preferably be refrigerated (https://www.transfusionguidelines.org/red-book/chapter-21-tissue-banking-tissue-retrieval-and-processing/21-2-retrieval). For other tissues, it is also recommended that the body should be refrigerated within an ideal window of 6 hours after death and the procurement of tissues needs to be completed within 24-48 hours. If not refrigerated, the tissues (excluding corneas) will then need to be retrieved within 12 hours of death due to the risk of tissue contamination. As the responsibility of certifying death often rests on the junior doctors, they play a vital role in determining the promptness in death certification, body refrigeration and subsequent eye retrieval, which has an important influence on the quality of the donor corneas.^11, 12^

We observed that the amount of ophthalmology teaching provided during the undergraduate training was low. Therefore, it would be difficult or impractical to incorporate teaching on corneal donation in undergraduate training. Moreover, depending on the clinical rotation during the Foundation Year programme, many doctors may never be involved in the discussion of organ/tissue donations. In view of these issues, training on corneal (and tissue) donation may be best targeted during postgraduate training, particularly in training rotations that usually deal with end-of-life care such as intensive care and oncology specialties. A potential strategy could be to incorporate a short mandatory induction training course at the start of these training rotations to improve the knowledge of corneal and tissue donation. As discussion of corneal and tissue donation with the family is relatively low among the junior doctors, we suggest that training of junior doctors should focus more on raising the awareness of the time limits and earlier death certification and notification of the relevant eye donation/retrieval personnel, instead of improving the knowledge of the actual contraindications of corneal and tissue donation.

Studies have shown that an opt-out system does not always translate to improved organ/tissue donation rate.^13^ One of the potential barriers could be attributed to the misperception of the opt-out system as shown in our survey. We observed that 61% of the junior doctors were incorrect about the fact that donation can proceed in the event of family refusal under the new opt-out system. Such misperception may affect the rate of donation as the junior doctors would not take the initiatives to discuss about organ/tissue donations, assuming that presumed consent automatically translates to donation.

One of the study limitations was that this survey only included the junior doctors working in the East Midlands, UK. However, these junior doctors were likely to have graduated from different medical schools, as reflected by the difference in the amount of undergraduate ophthalmology training received. It would be useful to survey the junior doctors working in other regions of the UK to enable a more generalised assessment of the knowledge and postgraduate training in corneal donation in the future. The reason we chose to survey junior doctors because they are the group of doctors who are most commonly involved in the initial process of death certification, which triggers the notification of the eye donation team. Secondly, the accuracy of the collected data relied on the honesty of the participants as this study was performed as an online survey. However, the relatively low number of correct responses provided by the respondents suggests that these were likely the true responses of the junior doctors.

In conclusion, our survey highlights the lack of knowledge of corneal donation and the new opt-out system among the junior doctors in the UK. Given the persistent shortage of donor corneas and the recent impact of COVID-19 on corneal donation, further targeted postgraduate training, particularly in specialties that deal with end-of-life care, could potentially enhance the corneal donation rate in the future.

## Data Availability

All the data have been provided in this manuscript.

## CONTRIBUTORSHIP STATEMENT

Study conceptualisation and design: BP and DSJT

Data collection: BP, OATL, PF, NEN, and all collaborators

Date interpretation: BP, EB, DGS, HSD and DSJT

Manuscript drafting: BP and DSJT

Critical revision of the manuscript: OATL, PF, NEN, EB, DGS and HSG

Final approval of the manuscript: All authors.

Overall supervision of the study: DSJT

## Acknowledgement

None

## FIGURES AND SUPPLEMENTARY DOCUMENTS

**Supplementary Table 1.** Factors that may influence the knowledge of corneal and tissue donation among the junior doctors in the UK. For analytic purpose, junior doctors were divided into two groups based on the number of correct answers (a total of 13 questions). Group A refers to those with less than 50% correct response (0-6 correct answers) and Group B refers to those with more than 50% correct response (7-13 correct answers).

| Parameters | Group A<br>Total N = 117;<br>N (%) | Group B<br>Total N = 26;<br>N (%) | P-value |
| --- | --- | --- | --- |
| Level of junior doctors |  |  | 0.36 |
| FY1 | 70 | 13 |  |
| FY2 or higher | 47 | 13 |  |
| Number of UG ophthalmology teaching, days | 10.8 ± 11.3 | 14.3 ± 15.2 | 0.19 |
| Previous ophthalmology training rotation during FYP |  |  | 0.71 |
| Yes | 19 (16.2) | 5 (19.2) |  |
| No | 98 (83.8) | 21 (80.8) |  |
| Previous discussion of corneal donation |  |  | 0.73 |
| Yes | 15 (12.8) | 4 (15.4) |  |
| No | 102 (87.2) | 22 (84.6) |  |
| Willingness to donate own corneas |  |  | 0.16 |
| Yes | 68 (58.1) | 19 (73.1) |  |
| No | 49 (41.9) | 7 (26.9) |  |

UG = Undergraduate; FYP = Foundation year programme
Continuous values are presented in mean ± standard deviation.

**Supplementary Figure 1.**
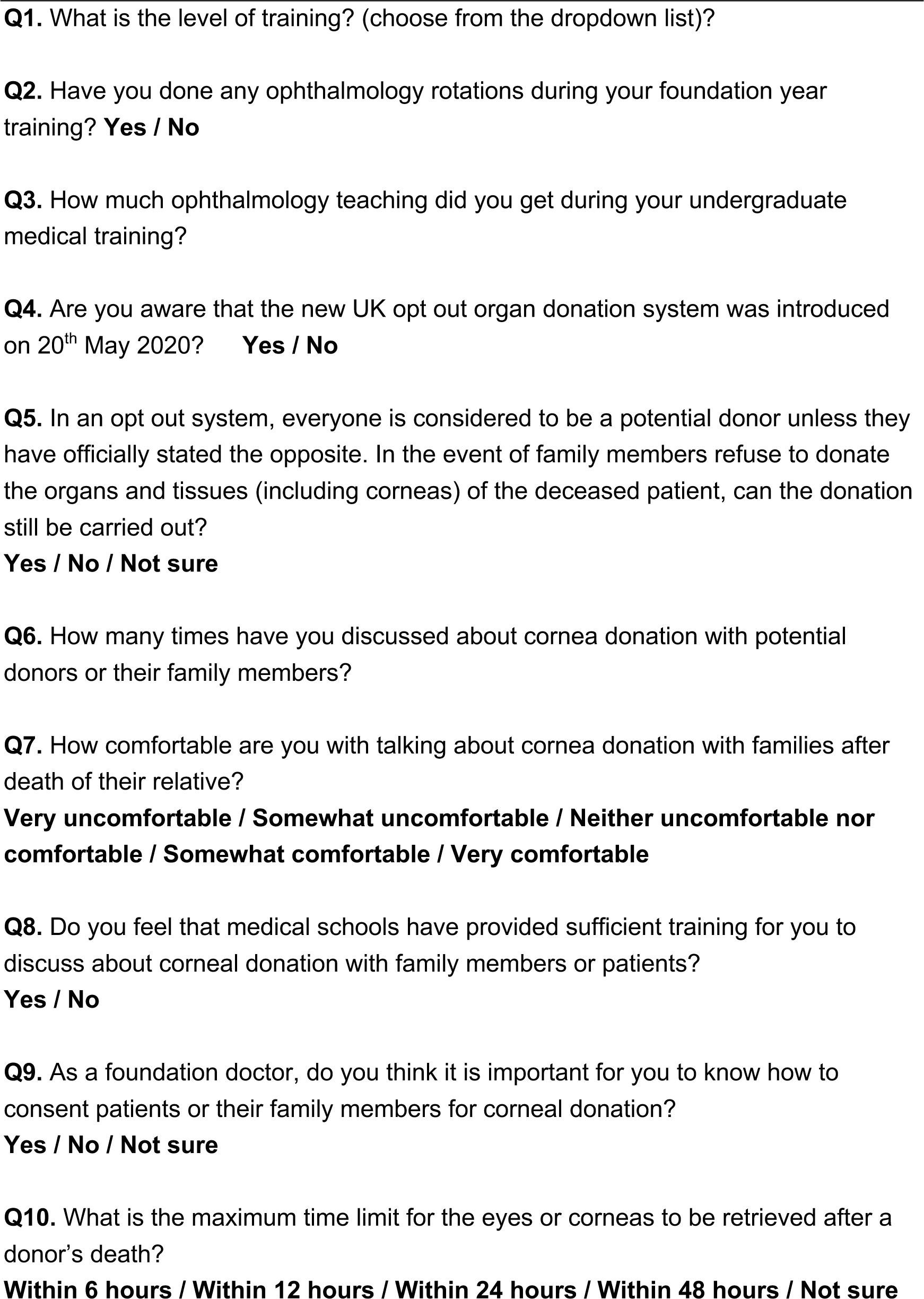
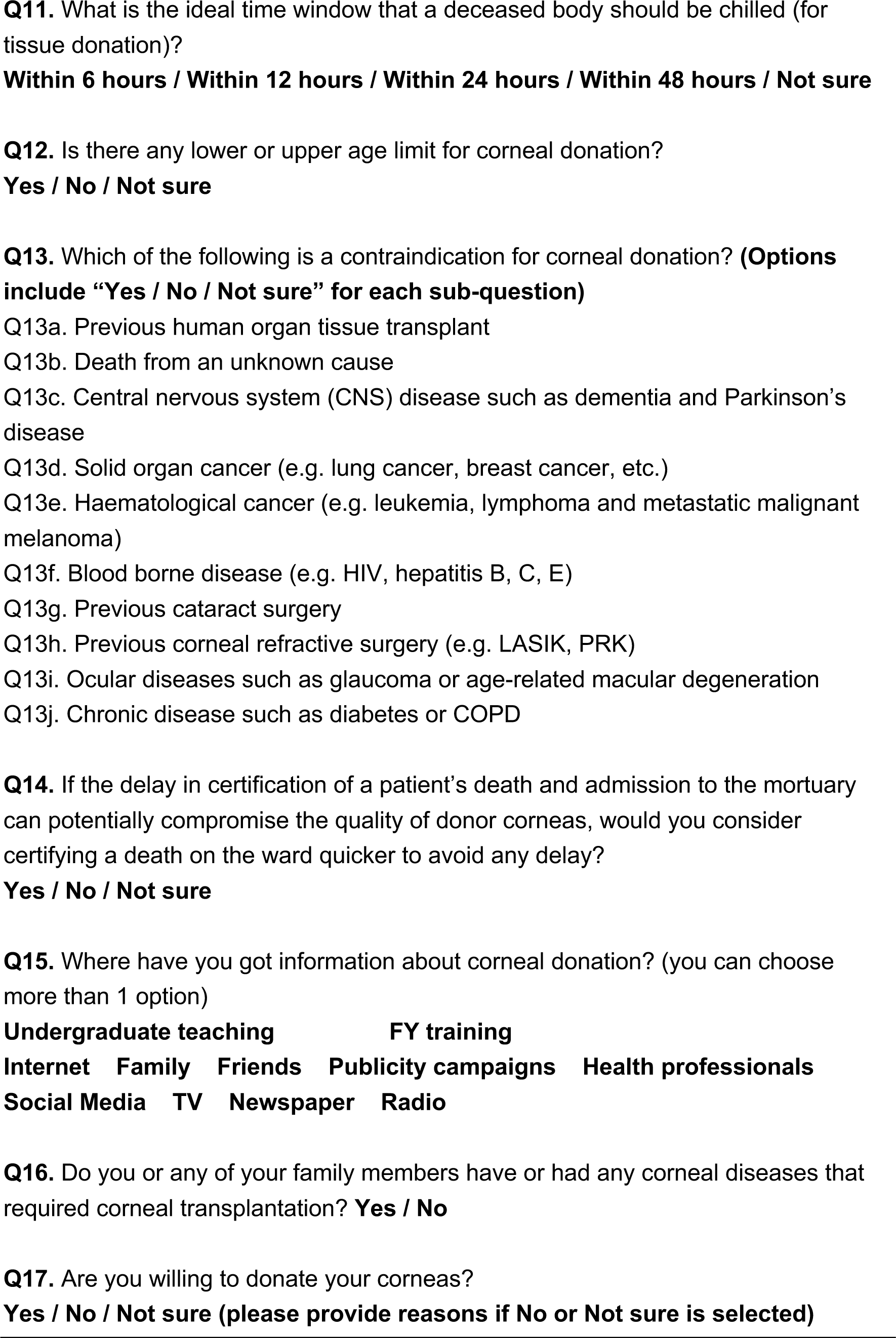
List of questions asked in the survey to evaluate the knowledge of cornea (and tissue) donation and the new opt-out system among junior doctors in the UK.

**Supplementary Figure 2.**
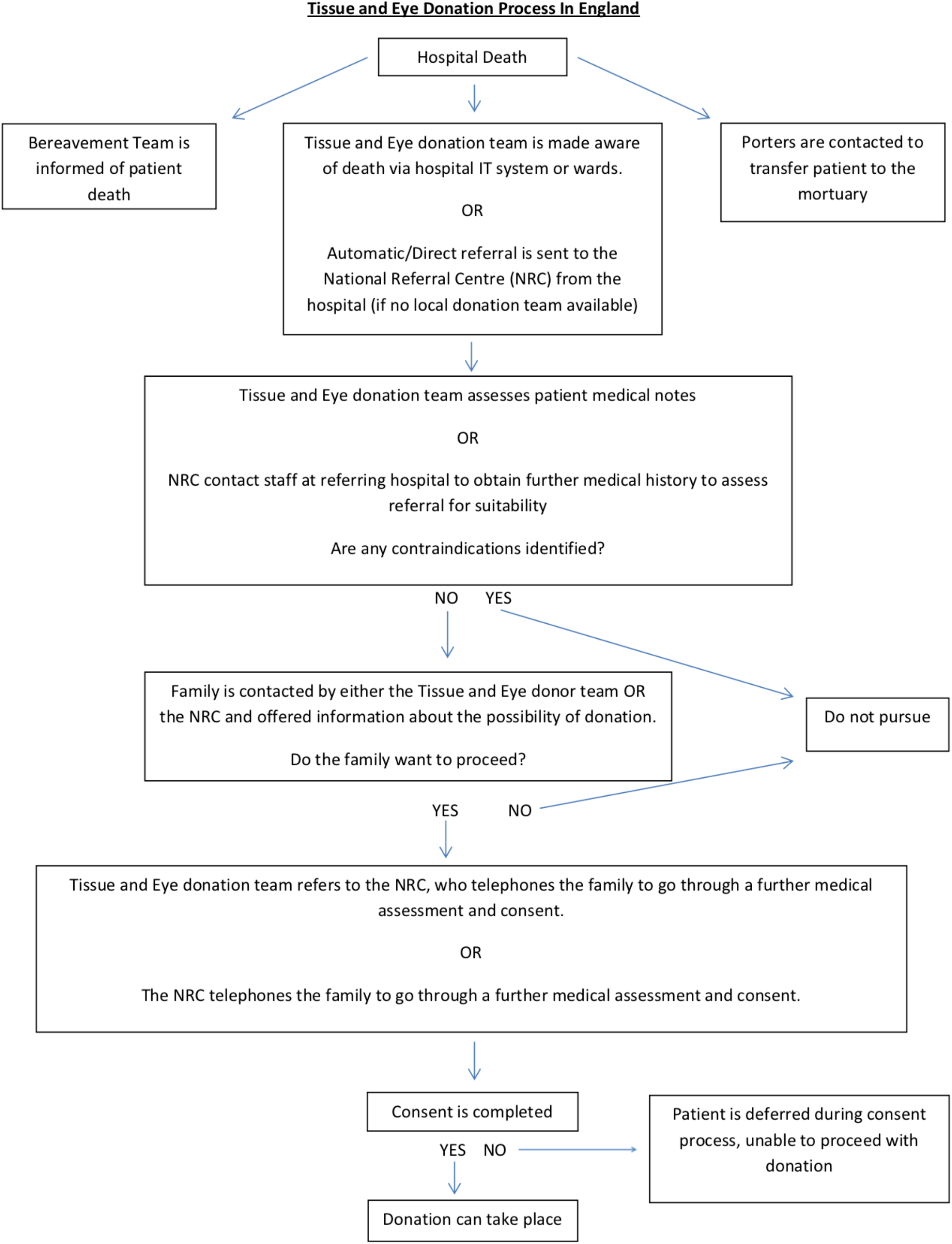
A summary of the eye donation process in England. This pathway highlights the important role of junior doctors, who are often the first point of contact in certifying hospital death and influencing the timeliness of body refrigeration in mortuary and notification of the eye / tissue donation team.

